# The role of serum brain injury biomarkers in individuals with a mild-to-moderate COVID infection and Long-COVID - results from the prospective population-based COVI-GAPP study

**DOI:** 10.1101/2023.02.15.23285972

**Authors:** Julia Telser, Kirsten Grossmann, Ornella C Weideli, Dorothea Hillmann, Stefanie Aeschbacher, Niklas Wohlwend, Laura Velez, Jens Kuhle, Aleksandra Maleska, Pascal Benkert, Corina Risch, David Conen, Martin Risch, Lorenz Risch

## Abstract

**Background:** During and after mild (no hospitalization) or moderate (hospitalization without ICU) SARS-CoV-2 infections, a wide range of symptoms, including neurological disorders have been reported. It is, however, unknown if these neurological symptoms are associated with brain injury and whether brain injury and related symptoms also emerge in patients suffering from Long-COVID. Neuronal biomarkers such as serum neurofilament light chain and glial fibrillary acidic protein can be used to elucidate neuro-axonal and astroglial injuries. We therefore investigated whether these biomarkers are associated with the COVID-19 infection status (mild-to-moderate), the associated symptoms and Long-COVID.

**Methods:** From 146 individuals of the general population with a post-acute, mild-to-moderate SARS-CoV-2 infection, serum neurofilament light chain (sNfL; marker of intra-axonal neuronal injury) and serum glial fibrillary acidic protein (sGFAP; marker of astrocytic activation/injury) were measured. Samples were taken before, during and after (five and ten months) a SARS-CoV-2 infection. Individual symptoms and Long-COVID status were assessed using questionnaires.

**Results:** Neurological symptoms were described for individuals after a mild and moderate COVID-19 infection, however, serum markers of brain injury (sNfL/sGFAP) did not change after an infection (sNfL: *P* = 0.74; sGFAP: *P* = 0.24) and were not associated with headache (*P* = 0.51), fatigue (*P* = 0.93), anosmia (*P* = 0.77) and ageusia (*P* = 0.47). In participants with Long-COVID, sGFAP (*P* = 0.038), but not sNfL (*P* = 0.58) significantly increased but was not associated with neurological symptoms.

**Conclusion:** Neurological symptoms in individuals after a mild-to-moderate SARS-CoV-2 infection with and without Long-COVID were not associated with brain injury, although there was some astroglial injury observed in Long-COVID patients.

**Funding:** The COVI-GAPP study received grants from the Innovative Medicines Initiative (IMI grant agreement number 101005177), the Princely House of Liechtenstein, the government of the Principality of Liechtenstein, and the Hanela Foundation (Switzerland). None of the funders played a role in the study design, data collection, data analysis, data interpretation, writing of the report, or decision to publish.

## Introduction

During the severe acute respiratory syndrome coronavirus 2 (SARS-CoV-2) pandemic, mild to severe neurological complications have been reported (Whittaker, Anson, and Harky 2020; Chou et al. 2021). Such neurological symptoms are often reported during the acute phase of the infection (Helms et al. 2020), but there is increasing evidence that they may persist for months, unrelated to the infection’s initial severity (Rogers et al. 2020). This lasting symptom burden for more than two months in COVID-19 patients can be linked to multiple organ systems and has led to the description of the post-COVID syndrome, also known as Long-COVID (Soriano et al. 2022).

Ultrasensitive Single Molecular Array (Simoa) assays allow the detection of diverse cerebrovascular injury in blood samples of acute and post-acute COVID-19 patients with high accuracy (Masvekar et al. 2022). One of those brain injury biomarkers is glial fibrillary acidic protein (GFAP) that is mainly expressed in astrocytes and regulates the function of these cells (Chmielewska et al. 2018). Astrocytic damage or activation can be indicated by increased GFAP levels in blood (Chmielewska et al. 2018). Other biomarkers are neurofilaments, exclusively expressed in neurons of the central and peripheral nervous system (Khalil et al. 2018; Yuan and Nixon 2021). Of the three neurofilament subunits, neurofilament light chain (NfL) has the lowest molecular weight and can diffuse from parenchyma to cerebrospinal fluid (CSF) and blood (Masvekar et al. 2022), making it a useful biomarker for neuro-axonal injury (Masvekar et al. 2022).

Recently, elevated levels of GFAP and NfL where found during the acute phase in blood samples of patients with severe (hospitalization with intensive care unit (ICU)) SARS-CoV-2 infections (Frontera et al. 2022; Aamodt et al. 2021; Prudencio et al. 2021; Cooper et al. 2020; De Lorenzo et al. 2021). However, despite the large number of patients with mild-to-moderate SARS-CoV-2 symptoms (Parasher 2021), only few studies investigated blood markers of brain injury in these groups during or even after resolution of the acute phase of the infection. One of these studies showed increased serum NfL (sNfL) and serum GFAP (sGFAP) levels in non-hospitalized adolescents (Havdal et al. 2022), while another study reported increased sNfL levels in adult health-care workers with mild-to-moderate symptoms (Ameres et al. 2020). To this date it is, however, still poorly understood whether the increase of the investigated biomarkers is associated with neurological symptoms in this groups. Therefore, to detect possible associations of sGFAP, sNfL and neurological symptoms and to assess whether biomarkers longitudinally change over time, we performed a follow-up study of patients after a mild-to-moderate SARS-CoV-2 infections. Also, we investigated whether sGFAP and sNfL change in Long-COVID patients after a mild-to-moderate infection, and whether these biomarkers are associated with Long-COVID symptoms.

## Methods

### Study design and participants

All participants from the population-based prospective cohort study GAPP (genetic and phenotypic determinants of blood pressure and other cardiovascular risk factors n = 2170) (Conen et al. 2013) were invited to participate in the sub-study COVI-GAPP. The COVI-GAPP study was initiated to investigate the use of a sensor bracelet (Ava-bracelet) to identify pre-symptomatic SARS-CoV-2 infections and detect infection-related physiological changes (Risch et al. 2022). Over the study period from 2020 – 2022, COVI-GAPP participants (n = 1144) were invited four times for blood collections at the study center in Vaduz, Liechtenstein.

For the current study, a post-acute SARS-CoV-2 infection was diagnosed with positive antibody results against the nucleocapsid (N) antigen of the SARS-CoV-2 virus. Only SARS-CoV-2 unvaccinated participants at the time of infection with a negative (control) and a minimum of one positive antibody-value against the SARS-CoV-2 N-antigen indicating seroconversion due to infection were included. From these, two participants were excluded due to missing baseline antibody values against the SARS-CoV-2 nucleocapsid (N) antigen, and seven participants were excluded due to insufficient sample volume, resulting in 146 participants with confirmed infection (**Figure 1**). From those, 88 participants had four tests (one negative, three positive), 30 participants had three tests (one negative, two positive) and 28 participants had two tests (one negative, one positive; **Figure 1**). Three participants could not be reached for clinical follow-up information, leading to 143 symptom queries for the respective analyses.

**Fig. 1:**
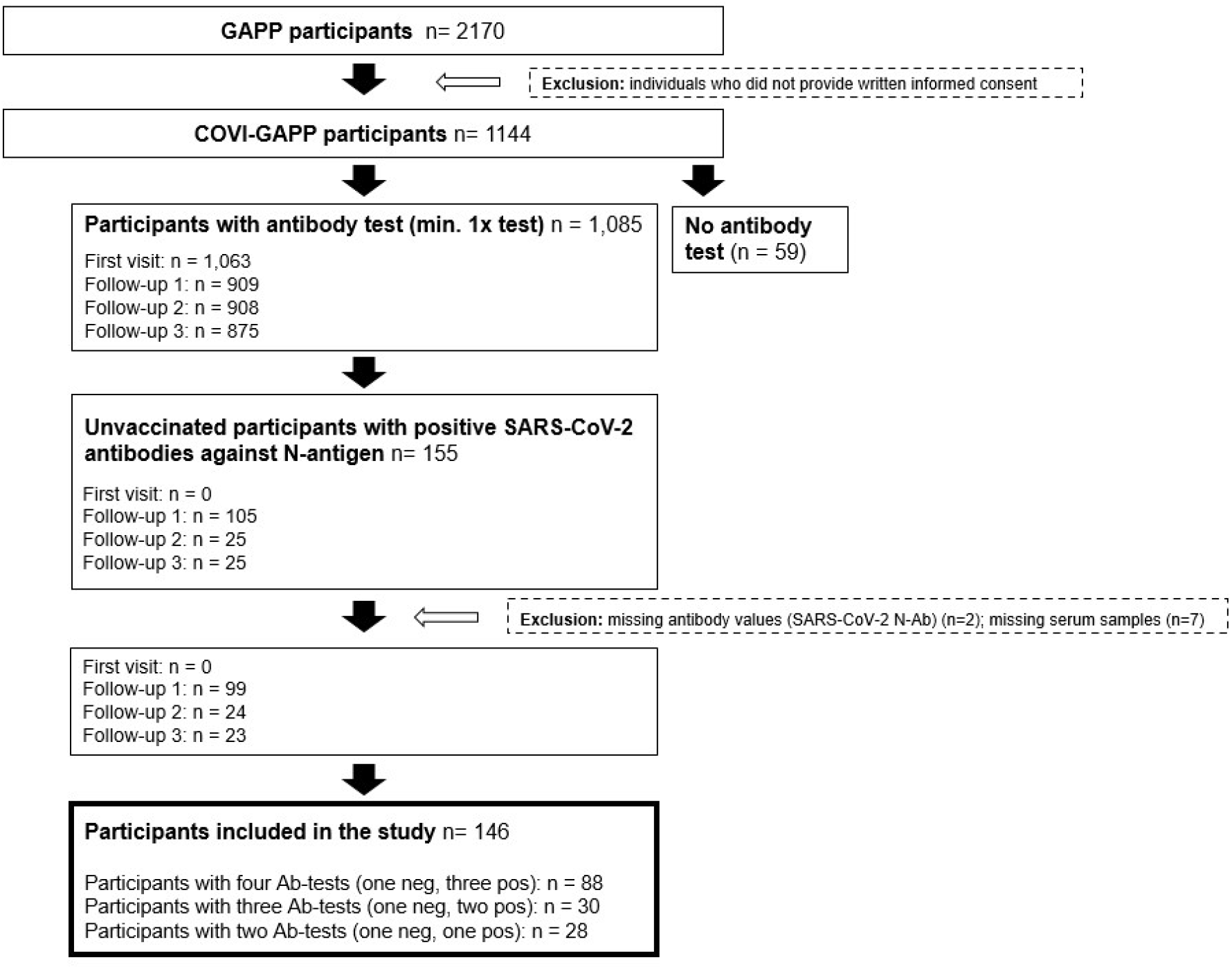
Study flow chart. From 2170 GAPP participants, 1144 participants were enrolled in the COVI-GAPP study. A total of 155 participants were unvaccinated and had a negative (control) and a minimum of one positive antibody-value against the SARS-CoV-2 nucleocapsid (N) antigen. After exclusion of nine participants due to missing data or serum samples, a total of 146 participants were included in the study. From those, 88 participants had four tests (one negative, three positive), 30 participants had three tests (one negative, two positive) and 28 participants had two tests (one negative, one positive).

From the 146 included participants, 133 participants had a mild infection, while 13 participants had a moderate infection. A mild infection is hereby defined as an infection that did not require hospitalization, while a moderate infection required hospitalization, but no intensive care unit (ICU). Thirty-nine participants with persistent symptoms of at least two months were considered as Long-COVID participants. From those participants, biomarker analysis (sNfL/sGFAP) was performed with the serum sample taken at the same time when participants firstly reported Long-COVID symptoms.

Informed written consent was obtained from each participant and the local ethics committee (KEK, Zürich, Switzerland) approved the study protocol (BASEC 2020-00786).

### Blood samples collection

Study-related blood collections only took place once participants were free of symptoms regarding active COVID-19 infection. At each visit, a venous blood sample was obtained from participants by trained study nurses in a standardized manner (Conen et al. 2013). Serum samples were kept at room temperature (RT) before SARS-CoV-2 antibodies (SARS-CoV-2-N-Ab and SARS-CoV-2-S1-Ab) analysis, which took place within 24 hours after blood collection. One aliquot was subsequently stored at -25°C before it entered a biobank for long term storage at -80°C.

### SARS-CoV-2 antibody measurements

SARS-CoV-2 antibody tests were assessed by Dr. Risch Ostschweiz AG, Buchs SG, Switzerland, an ISO 17025 accredited medical laboratory. Antibody levels were determined by electrochemiluminescence immunoassay (ECLIA) using the Elecsys® Anti-SARS-CoV-2 immunoassays (Roche Diagnostics, Rotkreuz, Switzerland) measured on a COBAS 6000. The Elecsys® Anti-SARS-CoV-2 S assay uses a recombinant protein representing the receptor-binding domain (RBD) of the spike (S) antigen or the nucleocapsid (N) antigen in a double-antigen sandwich assay format, which favors detection of high affinity pan-immunoglobulins directed against these SARS-CoV-2 antigens.

### Serum GFAP and serum NfL measurements

GFAP and NfL measures were performed at a median of 60 days (IQR: 32.0 to 77.5) after an acute infection. For GFAP and NfL analyses, the samples from the biobank were thawed, vortexed and aliquoted. Thereafter, they were frozen and shipped on dry ice to the University Hospital Basel, Switzerland for serum glial fibrillary acidic protein (sGFAP) and serum neurofilament light chain (sNfL) analysis (Khalil et al. 2020). sGFAP and sNfL measurements were performed with the commercially available Simoa Human Neurology 2-Plex B assay (N2PB, Item 103520) from Quanterix (Quanterix, Billerica, MA, USA) on the HD-X Simoa platform. Samples were analyzed in duplicate determination according to the manufacturer’s instructions.

All samples were analyzed by technicians blinded to the SARS-CoV-2 antibody values and health status of the participants. All sample concentrations were higher than the concentration of the lowest calibrator and lower than the concentrations of the highest calibrator. For GFAP, the mean inter-assay coefficient of variation (CV) of internal QCs was 9.7% (low), 10.6% (medium) and 9.0% (high) and for NfL 4.3% (low), 1.5% (medium) and 9.9% (high). For GFAP, the mean intra-assay CV from duplicate determination was 2.8% and for NfL, the mean intra-assay CV was 5%. Intra-assay coefficients of variation were below 15% for all analyses.

### Questionnaires

At the follow-up blood collections (five months and ten months post-infection), participants were asked to complete a written questionnaire, providing information about vaccination and infection status, and the duration of persistent symptoms (Long-COVID symptoms).

If participants had any symptoms during the study period, they were encouraged to visit the Liechtenstein National Testing Facility for reverse transcription-polymerase chain reaction (RT-PCR) testing, which was performed with either the COBAS 6800 platform (Roche Diagnostics, Rotkreuz, Switzerland) or the TaqPath assay on a QuantStudio 5 platform (Thermo Fisher Scientific, Allschwil, Switzerland) (Thiel et al. 2020; Goncalves Cabecinhas et al. 2021; Chen et al. 2021). Positively tested participants (PCR and antibody tests, or only antibody tests) were subsequently contacted by the study team and asked to report their symptoms (fever, fever degree, chills, cough, sniff, dyspnea, anosmia, ageusia, pressure in the chest, sore throat, muscle pain, headache, fatigue, general feeling of illness, diarrhea, sickness, vomiting) and hospitalization status by a standardized questionnaire commissioned by telephone interview (**Figure 2**).

**Fig. 2:**
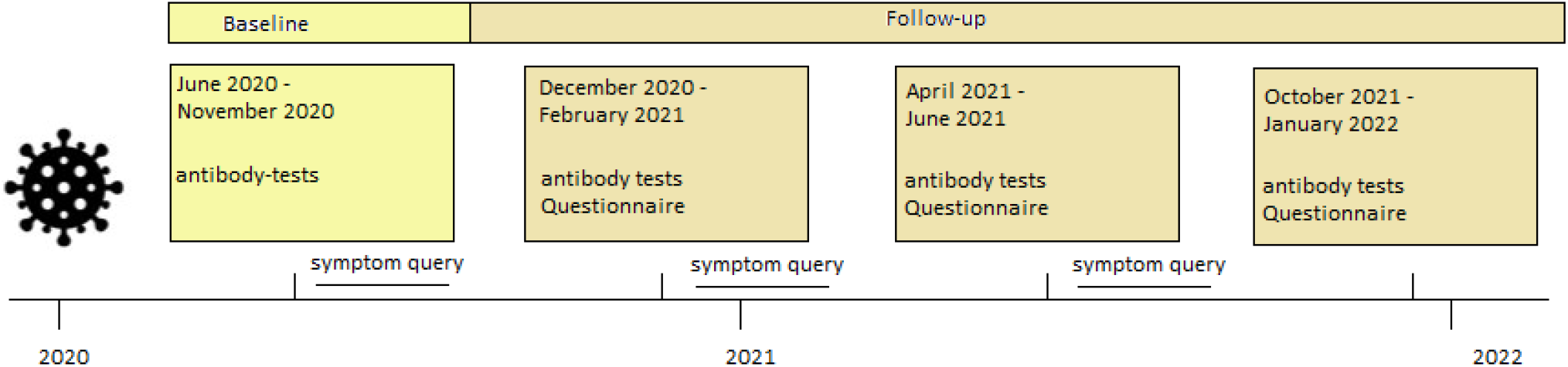
Time flow of data collection. Timeline of data collection. Each participant was invited for four antibody tests: An initial blood test (start June 2020, light yellow) and three follow-up blood tests (dark yellow). In each follow-up blood collection, a questionnaire was completed by the participants.

### Statistical analysis

We conducted the Mann-Whitney U test and chi-square test for demographics, stratified by infection severity (mild infection without hospitalization or moderate infection with hospitalization but no ICU). Distributions were assessed by visual inspection and outliers were detected using the Grubbs-right and Grubbs-left sided (alpha-level 0.05) test. The Wilcoxon test was used to assess biomarker (sNfL and sGFAP) differences between the SARS-CoV-2-N-Ab negative and SARS-Cov-2-N-Ab positive group. To assess differences between Long-COVID (symptoms for at least two months) vs no Long-COVID, the Mann-Whitney U test was used. To compare biomarker differences over time (follow-up of five months and ten months) we conducted the Friedman repeated measures test. To assess whether the reported symptoms are associated with brain injury biomarkers after a mild-to-moderate SARS-CoV-2 infection and with Long-COVID status, the Mann-Whitney U test was used. The Wilcoxon test was used to assess groupwise biomarker differences in participants with neurological symptoms before and after a SARS-CoV-2 infection. In addition, to assess whether sNfL and sGFAP is independently associated with the COVID status, a repeated measures ANOVA or a multivariable adjusted linear regression analysis was used with sNfL and sGFAP as dependent log-transformed variables and age, sex and COVID status as independent covariates. Outlier Analysis were conducted using MedCalc Version 20.027 and RStudio 2021.09.0 und R version 4.1.3.

## Results

### Demographics

Participant characteristics and clinical variables overall and stratified by infection severity are described in **Table 1**. Study participants were mainly infected by wild type, B.1.258 or B.1.1.7 variants, and B1.617.2. SARS-CoV-2 infections with Omicron were not observed within this period.

**Table 1:**
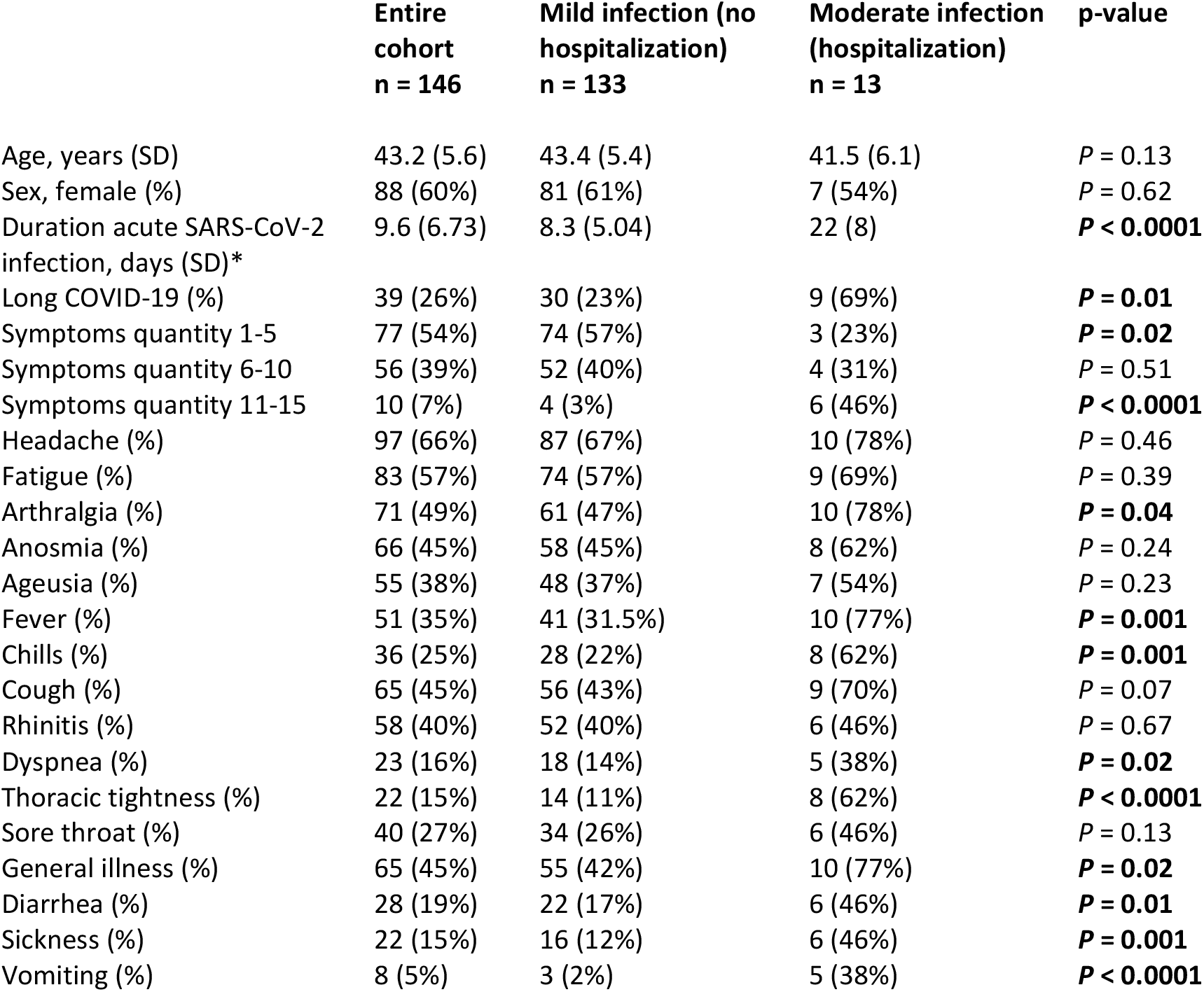
Demographics, serum biomarkers and symptoms. Data are shown as mean (SD) or n (%). Statistically significant difference was set at P < 0.05. * In the category “Duration acute SARS-CoV-2 infection”, four data are missing. Total: n = 142; mild infection: n = 129; moderate infection: n = 13. The Mann-Whitney U test and chi-square test was used to assess group differences.

The entire cohort (mildly and moderately infected participants combined) consisted of 146 unvaccinated participants at the time of infection, among which 57% got vaccinated (87.5% one shot and 12.5% two shots) during the average follow-up time of 16.7 months. Reinfection was reported in 1.4% of the cases of the entire cohort.

The median age of all participants was 43.2 years (SD = 5.6), and 88 (60%) participants were female. From the total of 146 individuals, 91% (133 participants) had a mild SARS-CoV-2 infection (without hospitalization) and 9% (13 participants) had a moderate SARS-CoV-2 infection (with hospitalization, but no ICU). There was no statistically significant difference between groups in term of age (*P* = 0.13) and sex (*P* = 0.62). The duration of the acute SARS-CoV-2 infection was significantly longer in participants with a moderate infection compared to those with a mild infection (*P* < 0.0001). Participants with a moderate infection were suffering more frequently from Long-COVID (*P* = 0.01).

The most reported symptoms for the entire cohort were headache (66%) and fatigue (57%). Participants with a mild infection reported less symptoms (1-5 symptoms), than participants with a moderate infection (11-15 symptoms) and most queried symptoms were reported more frequently in participants with a moderate infection. We found no statistical differences in the occurrence of neurological symptoms such as headache, fatigue, anosmia, and ageusia in patients with mild and moderate SARS-CoV-2 infections (**Table 1**).

sNfL was highly associated with participants age but not with participants sex (**Table 2**). For sGFAP, no association was found with either age or sex (**Table 2**). Since brain injury biomarkers are dependent on age, in all further analysis, the association between COVID-19 status and brain injury biomarkers was determined using a repeated measures ANOVA or multivariable adjusted linear regression analysis with sNfL or SGFAP as dependent log-transformed variables and age, sex and COVID-19 status as independent variables.

**Table 2.** Association of sNfL and sGFAP with age and sex. Multiple regression analysis of sNfL and sGFAP as dependent variables and age and sex as independent variables.

|  | *sNfL* |  | *sGFAP* |  |
| --- | --- | --- | --- | --- |
|  | Coefficient (Std. error) | p-value | Coefficient (Std. error) | p-value |
| Age | b1 = 0.11 (0.03) | ***P* = 0.002** | b1 = 0.54 (0.35) | *P* = 0.12 |
| Sex | b1 = 0.17 (0.43) | *P* = 0.78 | b1 = 5.27 (4.26) | *P* = 0.22 |

### Biomarkers of brain injury after a mild-to moderate SARS-CoV-2-infection

In the entire cohort, neither sNfL (*P* = 0.20) nor sGFAP (*P* = 0.12) levels significantly changed after an infection (median: 60 days, IQR: 32.0 to 77.5; **Fig. 3 A, B**).

**Figure 3.**
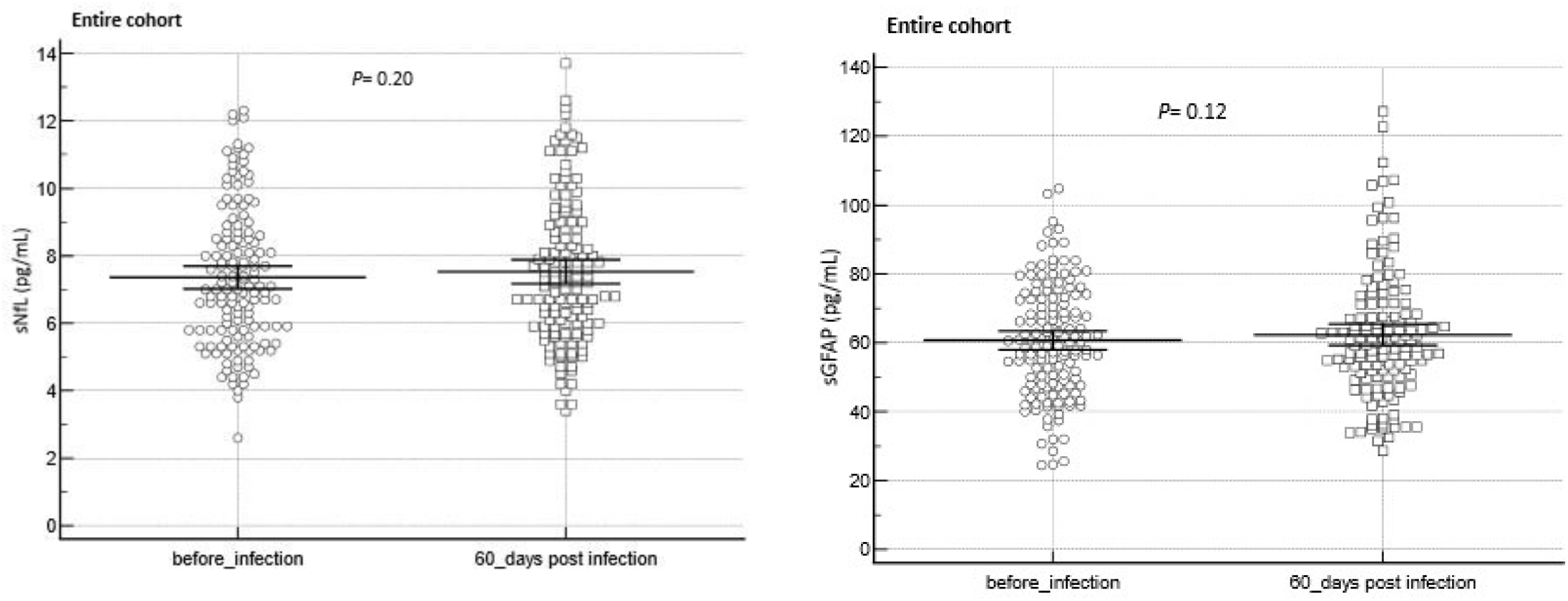
Brain injury biomarkers (sNfL/sGFAP) before and after a SARS-CoV-2 infection and at different clinical groups. **(A)** sNfL before and after infection (n = 142), **(B)** sGFAP before and after infection (n = 140). The Wilcoxon test was used to assess group differences before and after an infection. In the dot plot, complete cases were represented. The central horizontal line represents the mean with the 95% CI.

Over a follow-up period of ten months post-infection, sNfL (*P* = 0.74) and sGFAP (*P* = 0.24) levels didn’t change significantly over time **(Fig. 4 A, B)**.

**Figure 4.**
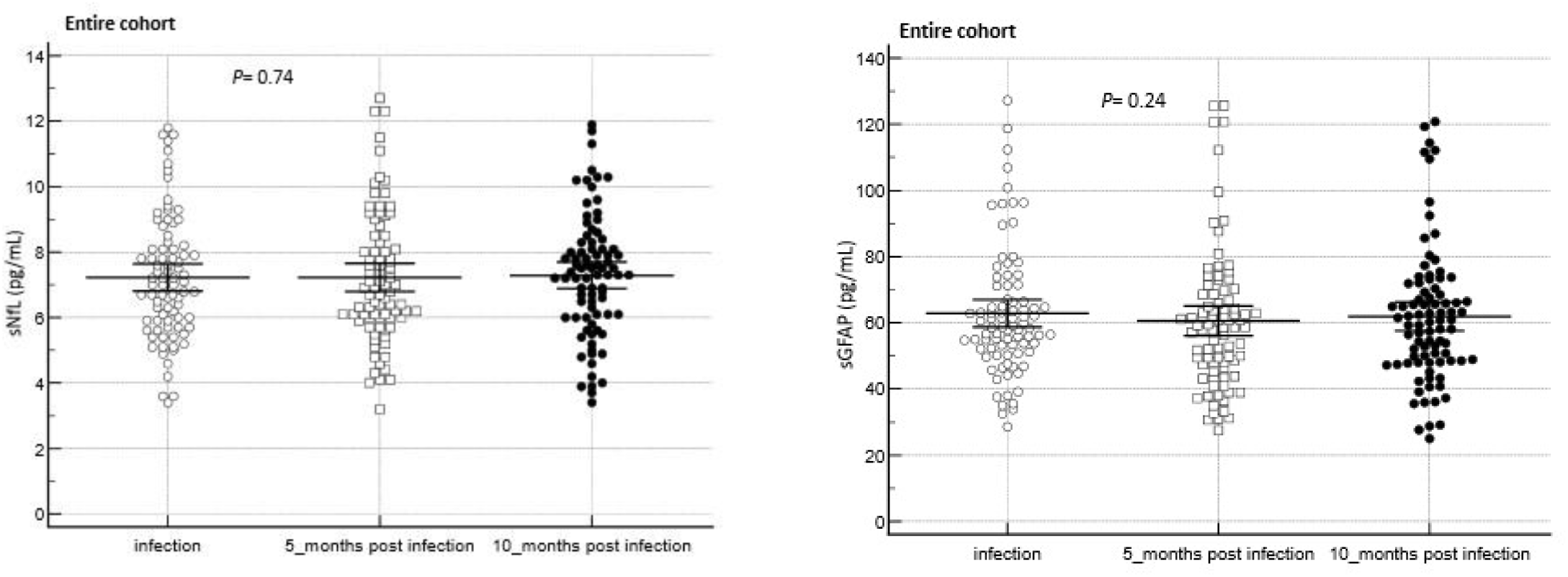
sNfL (n = 82) and sGFAP (n = 84) levels over time in the entire cohort **(A, B)**. To assess biomarker difference between the groups, the Friedmann test was used. In the dot plot, complete cases were represented. The central horizontal line represents the mean with the 95% CI.

In participants from the entire cohort suffering from neurological associated symptoms, sNfL and sGFAP levels did not change after an infection (**Table 3**).

**Table 3.** sNfL and sGFAP in the entire cohort with neurological associated symptoms such as headache (sNfL n = 93, sGFAP n = 91), fatigue (sNfL n = 78, sGFAP n = 75), anosmia (sNfL n = 64, sGFAP n = 60) and ageusia (sNfL n = 54, sGFAP n = 50) prior to a SARS-CoV2 infection and during the infection period. The Wilcoxon test was used to assess group differences.

| *symptoms* | *sNfL before infection vs after infection* |  |  | *sGFAP before infection vs after infection* |  |  |
| --- | --- | --- | --- | --- | --- | --- |
|  | **Median** | **IQR** | **p-value** | **Median** | **IQR** | **p-value** |
| headache | 7.1 vs 7.2 | 5.7 to 8.7 vs 5.9 to 8.8 | P = 0.25 | 60.5 vs 60.4 | 47.6 to 72.2 vs 51.3 to 68.1 | P = 0.51 |
| fatigue | 7.2 vs 7.2 | 5.9 to 9.2 vs 5.8 to 8.5 | P = 0.84 | 61.4 vs 59.4 | 47.6 to 72.2 vs 51.1 to 67.2 | P = 0.93 |
| anosmia | 12.3 vs 12.6 | 5.8 to 9.0 vs 5.7 to 8.8 | P = 0.88 | 60.7 vs 58.7 | 49.4 to 72.7 vs 51.3 to 67.4 | P = 0.77 |
| ageusia | 7.2 vs 7.2 | 5.9 to 8.9 vs 5.6 to 8.1 | P = 0.73 | 61.1 vs 56.9 | 47.9 to 75.5 vs 46.9 to 66.3 | P = 0.47 |

Although reported frequently during a SARS-CoV-2 infection, the occurrence of neurological associated symptoms such as headache, fatigue, ageusia and anosmia was not associated with sNfL or sGFAP levels in the entire cohort (**Table 4**).

**Table 4.** sNfL and sGFAP (median and IQR) in participants with vs without headache (sNfL n = 95 vs 46, sGFAP n = 90 vs 45), fatigue (sNfL n = 81 vs 60, sGFAP n = 78 vs 60), anosmia (sNfL n = 65 vs 76, sGFAP n = 61 vs 74) and ageusia (sNfL n = 54 vs 87, sGFAP n = 51 vs 85). The Mann-Whitney U Test was used to assess biomarker difference between the groups.

| *symptoms* | *sNfL* |  |  | *sGFAP* |  |  |
| --- | --- | --- | --- | --- | --- | --- |
|  | Median | IQR | p-value | Median | IQR | p-value |
| headache vs no headache | 7.2 vs 7.3 | 5.9 to 8.8 vs 6.0 to 9.0 | $P = 0.89$ | 60.4 vs 58.4 | 50.9 to 68.3 vs 50.0 to 72.2 | $P = 0.99$ |
| fatigue vs no fatigue | 7.2 vs 7.2 | 5.8 to 8.5 vs 6.2 to 9.0 | $P = 0.69$ | 59.7 vs 60.2 | 50.3 to 67.3 vs 50.1 to 75.9 | $P = 0.38$ |
| anosmia vs no anosmia | 7.4 vs 7.2 | 5.7 to 8.8 vs 6.5 to 8.9 | $P = 0.99$ | 58.5 vs 60.2 | 49.4 to 67.3 vs 50.1 to 71.5 | $P = 0.41$ |
| ageusia vs no ageusia | 7.2 vs 7.2 | 5.6 to 8.1 vs 6.1 to 9.1 | $P = 0.25$ | 56.2 vs 61.0 | 44.7 to 65.9 vs 50.7 to 71.5 | $P = 0.09$ |

### Biomarkers of brain injury in Long-COVID participants

No difference in sNfL levels were found between participants with Long-COVID (n = 38) and participants without Long-COVID (n = 91; *P* = 0.58)) in the entire cohort (**Fig. 5 A**). On the contrary, participants from the entire cohort with Long-COVID (n = 39) had significantly higher sGFAP levels compared to those participants without Long-COVID (n = 89; *P* = 0.038, **Fig. 5 B**).

**Figure 5.**
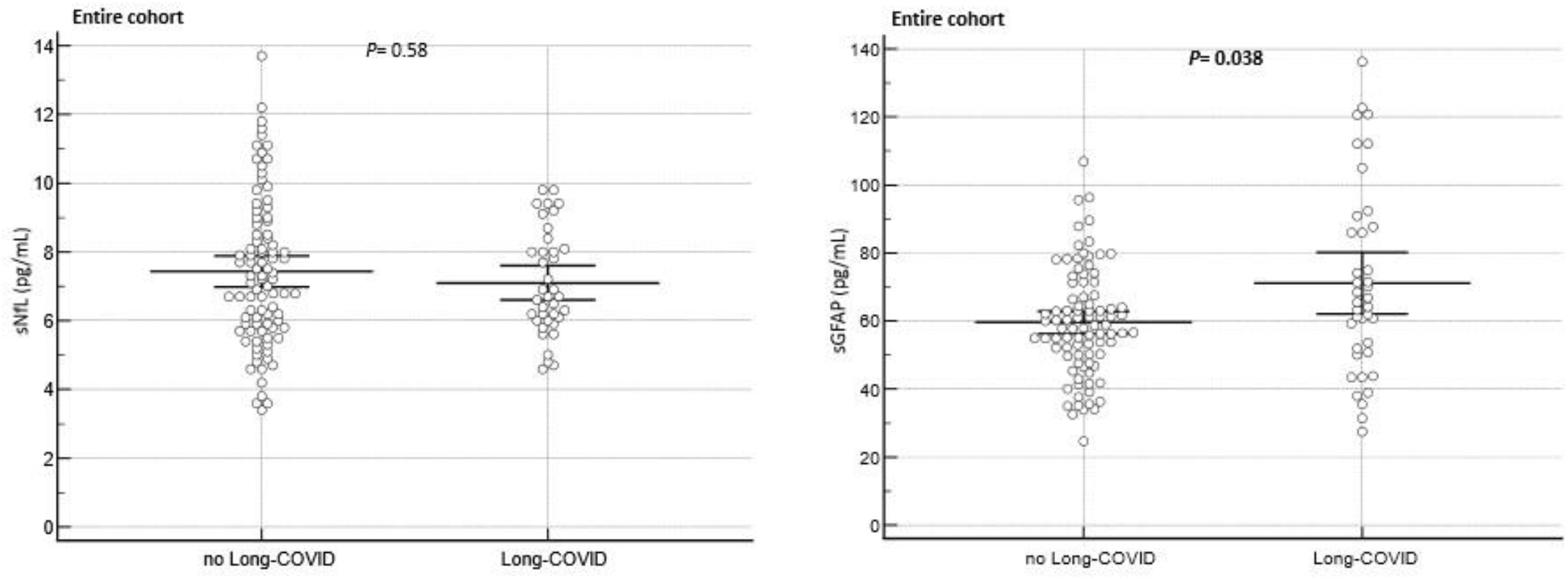
Brain injury biomarkers (sNfL/sGFAP) in patients with and without Long-COVID at different clinical groups. **(A)** sNfL in patients without vs with Long-COVID (n = 91 vs 38), **(B)** GFAP in patients without vs with Long-COVID (n = 89 vs 39). The Mann-Whitney U Test was used to assess biomarker difference between the groups. In the dot plot, complete cases were represented. The central horizontal line represents the mean with the 95% CI.

In Long-COVID participants suffering from neurological associated symptoms such as headache, fatigue anosmia and ageusia, sNfL and sGFAP levels not changed after an infection (**Table 5**).

**Table 5.**
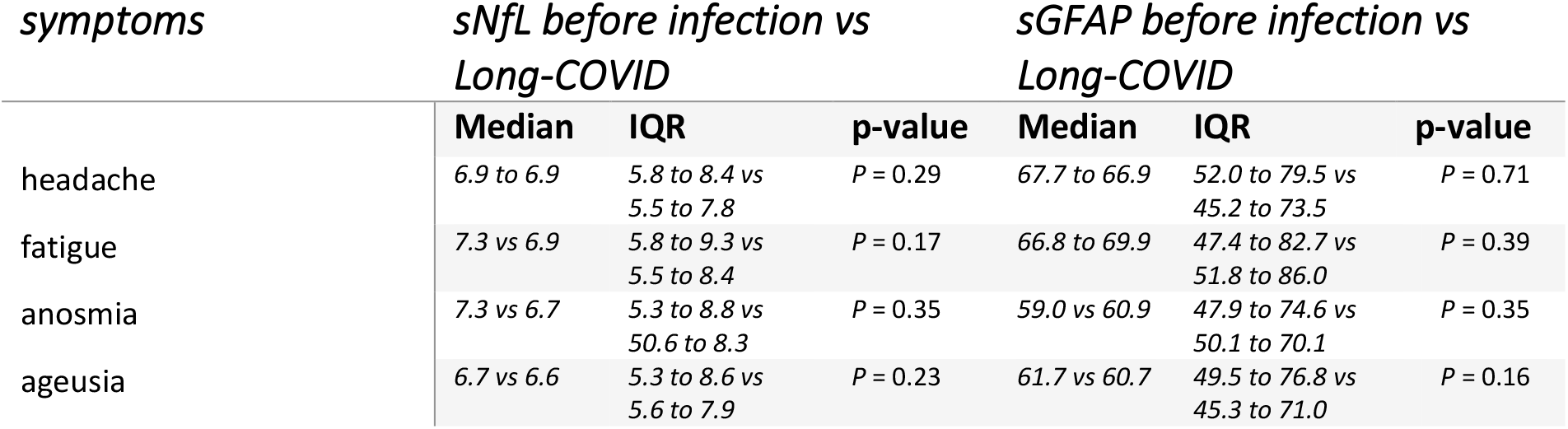
sNfL and sGFAP in Long-COVID patients with neurological associated symptoms such as headache (sNfL n = 16, sGFAP n = 15), fatigue (sNfL n = 21, sGFAP n = 14), anosmia (sNfL n = 19, sGFAP n = 18) and ageusia (sNfL n = 19, sGFAP n = 19) prior to a SARS-CoV2 infection and during the Long-COVID period. The Wilcoxon test was used to assess group differences.

Neurological associated symptoms such as headache, fatigue, ageusia and anosmia were not associated with sNfL or sGFAP in Long-COVID patients (**Table 6**).

**Table 6.**
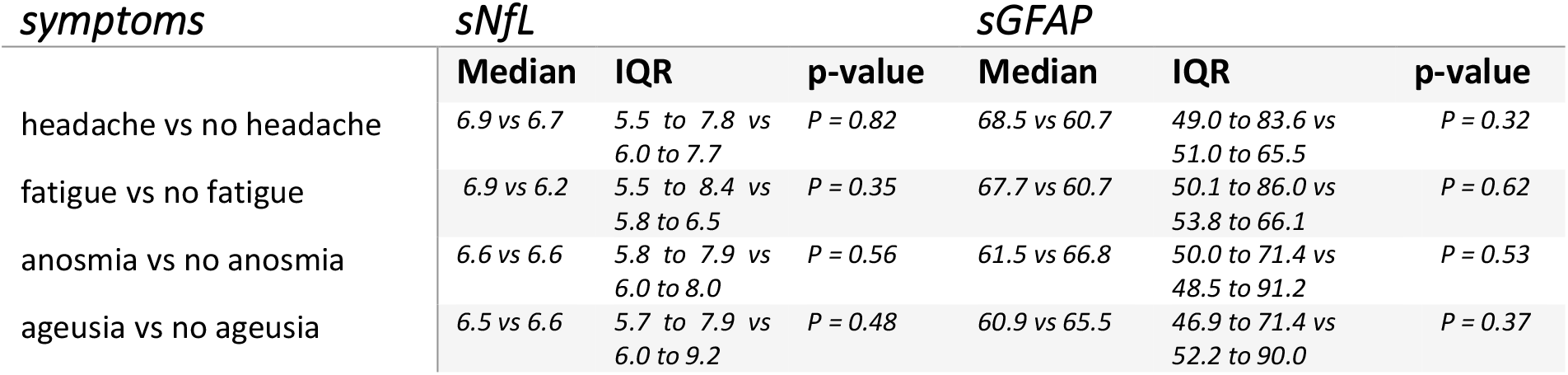
sNfL and sGFAP in the entire cohort in pariticipants with vs without headache (sNfL n = 16 vs 15, sGFAP n = 17 vs 16), fatigue (sNfL n = 21 vs 12, sGFAP n = 22 vs 11), anosmia (sNfL n = 20 vs 17, sGFAP n = 20 vs 17) and ageusia (sNfL n = 20 vs 14, sGFAP n = 20 vs 15). The Mann-Whitney U Test was used to assess biomarker difference between the groups.

## Discussion

In the present study we show that in participants with a moderate SARS-CoV-2 infection, the acute infection phase lasts longer, is accompanied by several symptoms and participants are affected more frequently of Long-COVID than mildly affected participants. Both sub-groups reported neurological associated symptoms, but the frequency did not vary between the groups. These reported neurological symptoms were not associated with serum markers of brain injury (sNfL/sGFAP) and in line with this, no association was found between the infection status and brain injury biomarkers, suggesting that COVID-19 infection and potentially associated neurological symptoms may not leave biochemical traces of neuro-axonal or astroglial damage after resolution in disease, at least not in peripheral blood samples.

Our results from non-ICU individuals after a mild-to-moderate infection suffering from Long-COVID indicate that sGFAP levels were related to Long-COVID. Our study therefore reported for the first time that Long-COVID status may be associated with brain injury. Interestingly, this seems to be a unique feature for sGFAP since no relation was found for sNfL in our cohort. This may suggest that the origin of some symptoms related to a Long-COVID differ from those symptoms coinciding with the acute or post-acute phase of the COVID-19 disease.

The lack of association of serum biomarkers of brain injury and infection status differs from previous studies, in which Ameres et al. (Ameres et al. 2020) found NfL to be elevated in adult health-care workers with a mild-to-moderate COVID infection and Havdal et al. (Havdal et al. 2022) found an increase in NfL and GFAP in non-hospitalized adolescents. This discrepant results to the latter study may be explained by the larger sample size, resulting in a more powered study and therefore, more likely to detect significant correlations. Moreover, our study investigated brain injury biomarkers in post-acute infection stages (60 days after infection), whereas Havdal et al. (Havdal et al. 2022) investigated biomarkers in the acute phase of the SARS-CoV-2 infection (not more than 28 days since the first day of symptoms or positive PCR test).

### Strengths and limitations

Strengths of our study include a well-defined group of unvaccinated individuals from the general population with a mild-to-moderate course of COVID infection. Moreover, our study included longitudinal data with a follow-up period of 10 months post-infection and participants with a mild-to-moderate COVID infection suffering from Long-COVID.

A weakness of the study is the small sample size of participants with a moderate infection not allowing subgroup analysis according to disease severity. In addition, symptoms and Long-COVID status, such as by definition, are self-reported and therefore vulnerable to recall bias. However, we think that these limitations do not invalidate our findings.

## Conclusion

Mild-to-moderate COVID-19 cases from the general population showed no association with brain injury biomarkers and neurological symptoms may not be a result of neuro-axonal or astroglial damage in those individuals. Individuals with a mild-to-moderate infection suffering from Long-COVID showed an increase in serum biomarker of astroglial injury but not neuro-axonal damage and therefore, our study reported for the first time that Long-COVID status may be associated with brain injury. To draw further conclusion, additional studies in individuals with mild-to-moderate COVID infection, with and without Long-COVID and sNfL/sGFAP are required.

## Data Availability

All data produced in the present study are available upon reasonable request to the authors

## Acknowledgement

We acknowledge the COVI-GAPP participants who enrolled in this study. In addition, we thank the study team in Vaduz, Liechtenstein and the different teams at the Dr. Risch Medical Laboratory in Buchs, Switzerland for their contribution to this study. Moreover, we thank the laboratory staff from the Neurologic Clinic and Policlinic, MS Center and Research Center for Clinical Neuroimmunology and Neuroscience Basel from the University Hospital Basel, Switzerland for contributing laboratory measurements.

## Competing interests

LR, and MR are key shareholders of the Dr Risch Medical Laboratory. DC has received speaker fees from DMS/Pfizer and Servier, as well as consulting fees from Roche Diagnostics and Trimedics, all outside of the current work. The other authors have no financial or personal conflicts of interest to declare.

## Notes

### Author Declarations

Informed written consent was obtained from each participant and the local ethics committee (KEK, Zuerich, Switzerland) approved the study protocol (BASEC 2020-00786).

